# Intraindividual variability in cognitive function in older people in the English Longitudinal Study of Ageing: modelling risk factors using a mixed-effects beta-binomial model

**DOI:** 10.1101/2022.06.01.22275869

**Authors:** Richard M.A. Parker, Kate Tilling, Graciela Muniz Terrera, Jessica K. Barrett

## Abstract

Cognitive functioning in older age has a huge impact on quality of life and physical and mental health. Whilst most research in cognition in older age has focussed on mean levels, there is some evidence that individuals with cognitive functioning that varies a lot around this may have different risk factors and outcomes to those with less variable functioning. Existing approaches to investigate such intraindividual variability (IIV) typically involve deriving a summary statistic for each person from residual error around a fitted mean. However, such methods ignore sampling variability, prohibit the exploration of associations with time-varying factors, and are biased by floor and ceiling effects. To address this, we fitted a mixed-effects location scale beta-binomial model to estimate average per-trial probability and IIV in a word recall test with the English Longitudinal Study of Aging (ELSA). After adjusting for mean performance, in an analysis of 9,873 individuals observed across 7 (mean: 3.4) waves we found IIV to be greater: at older ages; with lower education; in females; with more difficulties with activities of daily living; in later cohorts; and when interviewers recorded issues which may have affected the tests. Our study identifies groups with more varying cognitive performance, which has implications for their daily functioning and care. Further work is needed to identify the impact of this for future health outcomes.

---

Cognitive functioning in older people has profound implications for current and future health and wellbeing [1, 2]. In the absence of therapeutical cures for dementia, for example, changes in cognitive performance can aid the early identification of individuals at increased risk of developing the condition, paramount for the design and implementation of interventions that may delay the onset of faster deterioration [3, 4]. Traditionally, research in cognitive decline has focused on the study of individual differences and on the identification of risk factors for rate of mean change [5]. However, some have investigated inconsistency in performance [6]. For example, evidence is emerging that inconsistency across different cognitive tasks in a single occasion (cognitive dispersion) is a potential early marker of pathological changes in the brain and shows its association with critical outcomes [7-9]. In addition, intraindividual variability (IIV) in performance can be measured in the same task over shorter (same visit) or longer (visit-to-visit) periods of time. MacDonald, Li and Backman reviewed the empirical evidence linking IIV in cognitive performance with neural correlates and discussed possible mechanisms that may explain such associations [10]. With a focus on IIV in reaction time, Kochan *et al*. used data from the Sydney Memory and Ageing Study, a longitudinal study of older adults in Australia, and reported that greater IIV, but not greater mean reaction time, significantly predicted survival time after adjusting for sociodemographic factors, cardiovascular risk index and apolipoprotein ε4 status [11]. Similarly, Gamaldo *et al*. examined differences in IIV between impaired and unimpaired participants of the Baltimore Longitudinal Study of Aging, and showed that individuals who had received a diagnosis of dementia had greater variability in attention, executive function, language and semantic memory at least 5 years before the onset of cognitive impairment compared to individuals who remained free of dementia, demonstrating the potential role of IIV as an early indicator of pathological changes [12].

Despite the increasing interest in IIV, the analytical approaches commonly used to quantify it in longitudinal studies are limited. Some researchers have considered the average amount of deviation (residual error around a fitted mean) in an individual’s performance over time [13, 14]. However this does not adjust for sampling variability given the finite number of within-person observations, and the resulting individual-level summary statistic is not amenable to the exploration of associations of IIV with time-varying factors. Alternatively, Gamaldo *et al*. fitted multilevel (MLM) or growth curve models to repeated measurements of the outcome of interest, and then compared models which assume the residual IIV to be constant to models which allow it to depend on fixed effects, e.g. diagnostic status [12]. However, this assumes that people within each group have the same IIV. We have previously shown that in an MLM, the residual IIV can instead be assumed to contain systematic variation that can be explained, depending not only on fixed effects (as in Gamaldo *et al*. [12]), but on random effects as well, in mixed-effects location scale (MELS) models [15-17]. MELS models allow for the association of IIV with predictors which may be time-varying, or otherwise, to be investigated. They further allow for residual differences between people in their IIV to be estimated via random effects, and their association with the individual mean to be investigated via correlated random effects [18].

Whilst MELS models typically assume that, conditional on the random effects, the response variable is Normally distributed, this is likely to be violated with a bounded discrete outcome [19], where floor and ceiling effects can lead to underestimated IIV for people returning high, or low, mean scores. Under such circumstances, a beta-binomial model has been shown to improve statistical inference [20]. Beta-binomial models have a location (*p*) and scale (*θ*) parameter, each of which can be allowed to differ across fixed and random effects in an analogous manner to a MELS model. Given *θ* captures heterogeneity in the average per-trial probability, factors associated with IIV can thus be investigated.

Since the evidence on factors associated with differences in IIV over time is limited, we use a MELS beta-binomial model to investigate visit-to-visit IIV in a word recall test in the English Longitudinal Study of Ageing (ELSA). Our aim is to understand the factors associated with IIV in a test of episodic memory in older adults, using a modelling approach appropriate to this bounded discrete outcome.

## METHODS

### Cohort

Participants were from the English Longitudinal Study of Ageing (ELSA) [21], an ongoing panel study that contains a nationally representative sample of the English population aged 50 and over living in households, previously described [22, 23]. Interviews at baseline (2002–2003) were carried out with 11,391 individuals (5,186 men and 6,205 women); the overall response rate was 70% at the household level and 67% at the individual level. After the baseline interview, follow-up interviews took place at regular 2-year intervals in 2004–2005 (wave 2), 2006–2007 (wave 3), 2008–2009 (wave 4), 2010–2011 (wave 5), 2012–2013 (wave 6) and 2014-2015 (wave 7). Refresher samples were added at waves 3, 4, 6 and 7 to ensure the study remained representative of the target age group. Participants gave full informed consent to participate in the study. We restricted our sample to core participants responding to at least one wave when aged 65 years old or older, and to observations for which the participant did not report having Alzheimer’s disease or dementia, organic brain syndrome, senility or any other serious memory impairment.

### Cognitive function

Memory was measured using a 10-word recall test that has earlier been used in the Health and Retirement Study [24]. Participants were presented with a list of 10 words that were read out to them and asked to recall as many words as they could both immediately and, with no prior notice, five minutes later and after they had been asked to complete other survey questions. A total of four versions of the 10-word lists were available and were randomly allocated by computer. The number of correctly recalled words was used as a measure of memory (range: 0–20 words) adding the results from both the immediate and delayed recall tests.

### Covariates

Information on participants’ age, sex, education, and difficulties with Activities of Daily Living (ADLs) were recorded at each wave. In addition, the interviewer reported whether there were any factors which may have impaired the participants’ performance during the cognitive tests. Education was categorised into higher (college / university), secondary and no qualifications. For difficulties with ADLs, participants were asked if they had any difficulty dressing (including putting on shoes and socks), eating (including cutting up food), bathing and showering, getting in and out of bed and walking across a room, and a scale counting the number of items participants had difficulties with was derived from this. Interviewer-recorded factors which may have affected the cognitive tests included: the participant being blind or having poor eyesight, being deaf or having poor hearing, being too tired, illness or physical impairment, impaired concentration, being very nervous or anxious, having other mental impairment, an interruption or distraction, a noisy environment, problems with the testing computer, difficulty in understanding English, or any other factors. This was a binary variable, indicating whether there were no, or at least one, such issue recorded.

### Analytical approach

A mixed-effects location scale (MELS) beta-binomial model was fitted to repeated measurements of the longitudinal outcome, the word recall test score. The beta-binomial model assumes that each observed value of the outcome (each score out of 20, in our case) has an underlying, unobserved probability which is sampled from a beta distribution. The shape of the beta distribution from which these probabilities are drawn is defined by an average per-trial probability parameter *p* (a.k.a. *μ*) and a variability (or scale or dispersion) parameter *θ* (a.k.a. *φ* or *κ*) [25, 26]. When *θ* is 2, then every probability, from 0 to 1, is equally-likely. When *θ* < 2 then dispersion is greater and extreme probabilities near 0 and 1 become more likely than the mean, whilst when *θ* > 2 the distribution of probabilities becomes concentrated around the mean [25, 27]. Supplementary Figure S1 plots expected distributions of test scores given different values for these parameters.

Instead of directly modelling the probability for each observed count, the beta-binomial models this distribution of probabilities, via *p* and *θ*. Whilst a MELS model is typically a Gaussian model, we use the terminology here as we include both fixed (population) and random (individual) effects in (1) the linear predictor for *p* (the ‘location’ of the beta distribution), and (2) in the linear predictor for *θ* (the ‘scale’ of the beta distribution, where low estimated values of *θ* imply greater intraindividual variability (IIV) in task performance).

Covariates added to the linear predictor for *p* were age, cohort (the year of reaching age 65), sex, educational qualification, the number of ADLs with which the respondent reported difficulty, and whether the interviewer reported whether there were any factors which may have impaired the participants’ performance during the cognitive tests. These covariates were also included in the linear predictor for *θ*. For the linear predictor for *p*, any non-linearity in the association between age and the outcome was first assessed by fitting restricted cubic regression splines with different sets of knots as recommended by Harrell (2015) [28], and the best-fitting function of age was selected and fitted. Interactions of age with each of sex, educational qualifications, number of ADL difficulties, and issues potentially impairing test performance were also added, in turn, to the linear predictor for *p* to see if they improved model fit. Model fit was assessed via Pareto smoothed importance sampling leave-one-out (PSIS-LOO) cross-validation [29]. In addition, random effects were included in both the functions for *p* and *θ* to account for unobserved heterogeneity between individuals. In the predictor for *p*, a random intercept estimated the between-individual variability in *p* at the mean age, and a random slope estimated the between-individual variability of the effect of age (as a linear term fitted across the whole age range) on *p*. In the linear predictor for *θ*, a random intercept estimated the extent to which people differed in their IIV (specifically in how dispersed the beta distribution from which the underlying probability of test success was drawn). Random effects were assumed multivariate normally distributed, allowing for non-zero correlations between them.

The models were fitted using Bayesian estimation via MCMC methods in Stan (2.21.0), using the brms package (2.16.1) in R (4.1.0) [30-32]. Results are reported as means of posterior distributions and 95% credible intervals. See the Supplementary Materials for further details of these (and other sensitivity) analyses, and sample code.

## RESULTS

A total of 9,873 individuals were included in the final cohort (see Supplementary Figure S2). Of these, n = 2,202 (22.3%) were reported as dying during the study period, whilst the mortality status of 2,477 (25.1%) was reported as unknown at the final wave (wave 7), with the remaining 5,194 (52.6%) reported as alive at that wave. N = 396 (4.0%) of the final cohort were reported as having a memory problem in at least one survey (these, and subsequent, surveys for such participants were not included in the model). On average, participants included in the final cohort contributed data to 3.4 data collection waves, distributed across waves as follows: 1^st^ data collection wave: n = 5,283 (53.5%); 2^nd^: n = 4,566 (46.2%); 3^rd^: n = 4,149 (42.0%); 4^th^: n = 4,728 (47.9%); 5^th^: n = 4,811 (48.7%); 6^th^: n = 5,037 (51.0%); 7^th^: n = 4,917 (49.8%).

Table 1 shows the baseline characteristics of included individuals, including by whether they contribute data to every wave after becoming eligible or not. It indicates that those who did not contribute data to every wave after becoming eligible had, on average, a lower memory test score, were older, had lower educational qualifications, had more difficulties with ADLs, and had more issues recorded by the interviewer which may have affected the cognitive tests.

**Table 1.**
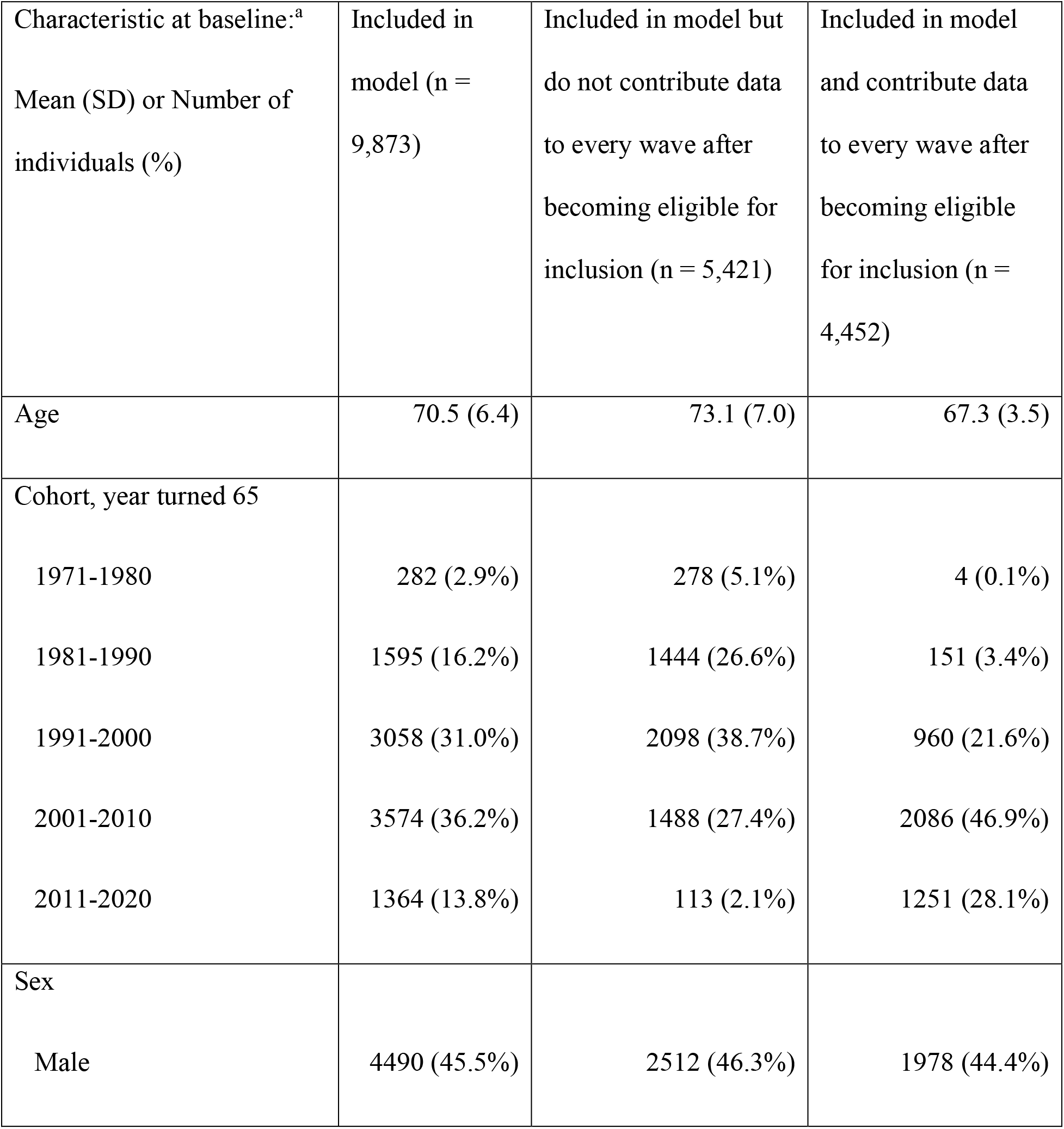
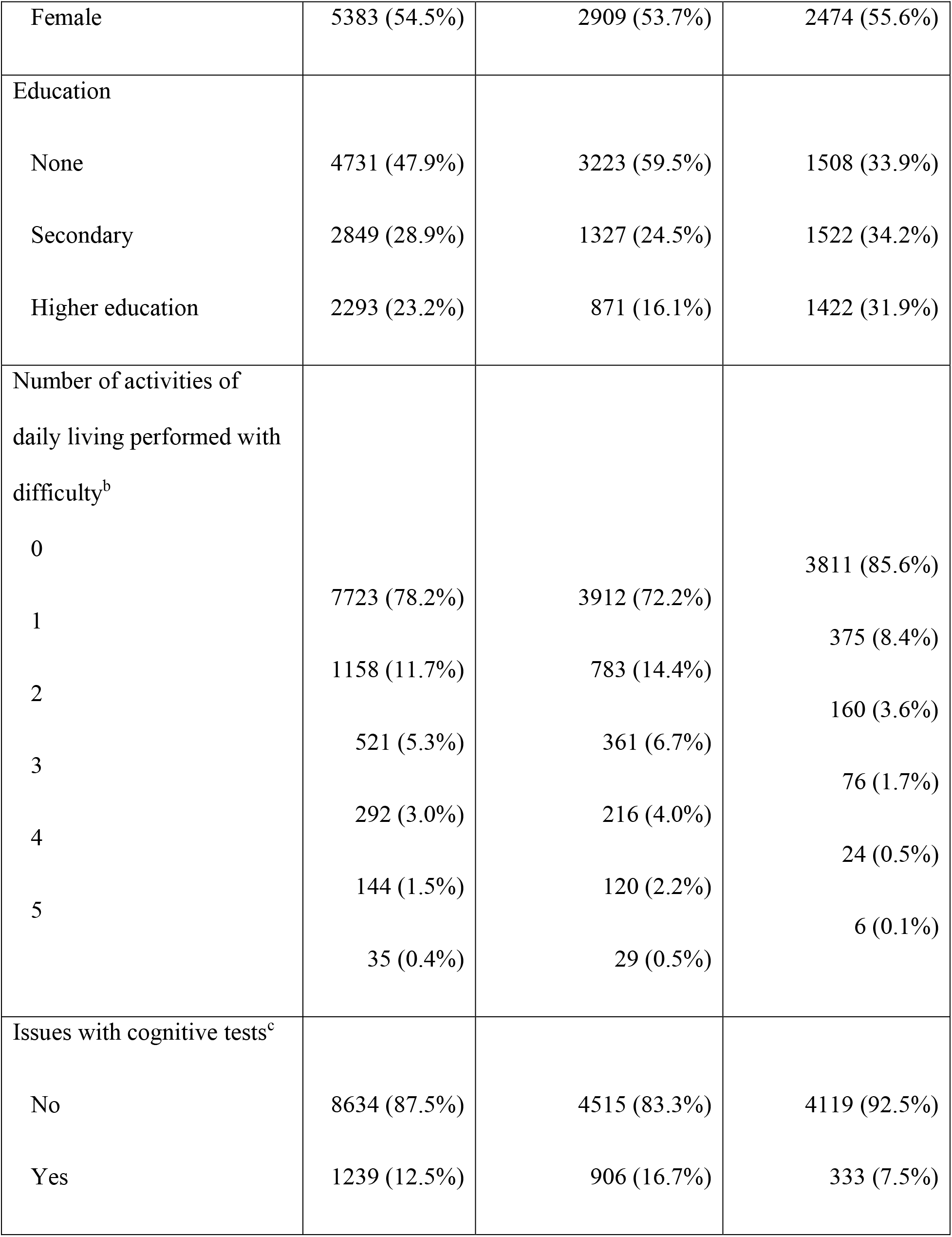
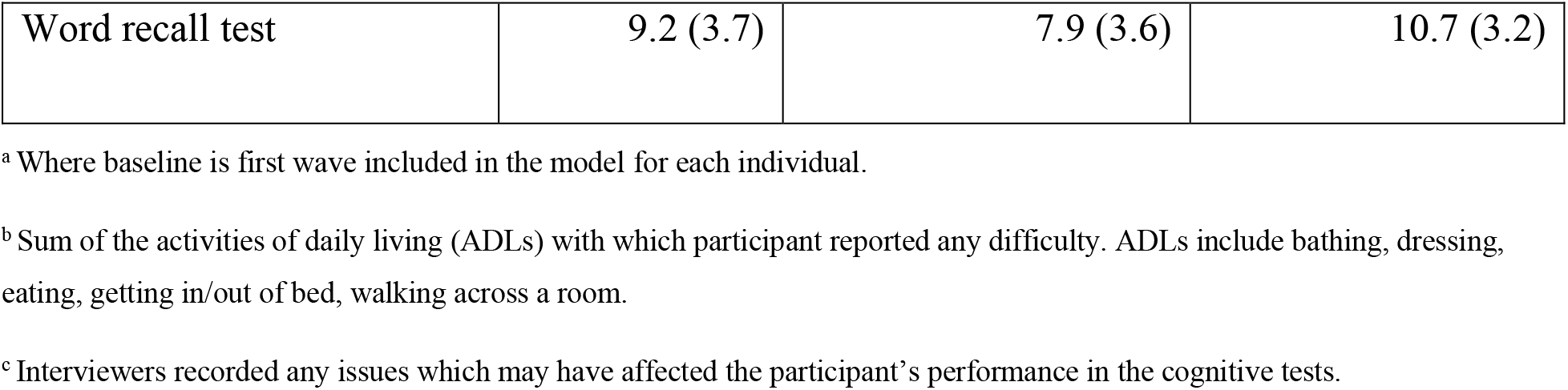
Summary statistics at baseline for individuals included in the model, further subdivided by whether they contribute data to every wave after becoming eligible for inclusion or not.

See Supplementary Materials for further summary statistics, including by exclusion status and drop-out status (Tables S1-S2), and also for all estimates from the models presented below (Table S3).

### Association of covariates with *p* (average per-trial probability)

A restricted cubic spline for age, with 4 knots points, was fitted in the fixed part of the linear model for *p* (see Supplementary Materials). Figure 1 presents the estimated average per-trial probability of providing a correct answer in the word recall test, across age. It indicates that, on average, this probability decreased with age – i.e. older participants were less likely to recall the words earlier presented – with the rate of this decline greater from around 80 years of age.

**Figure 1.**
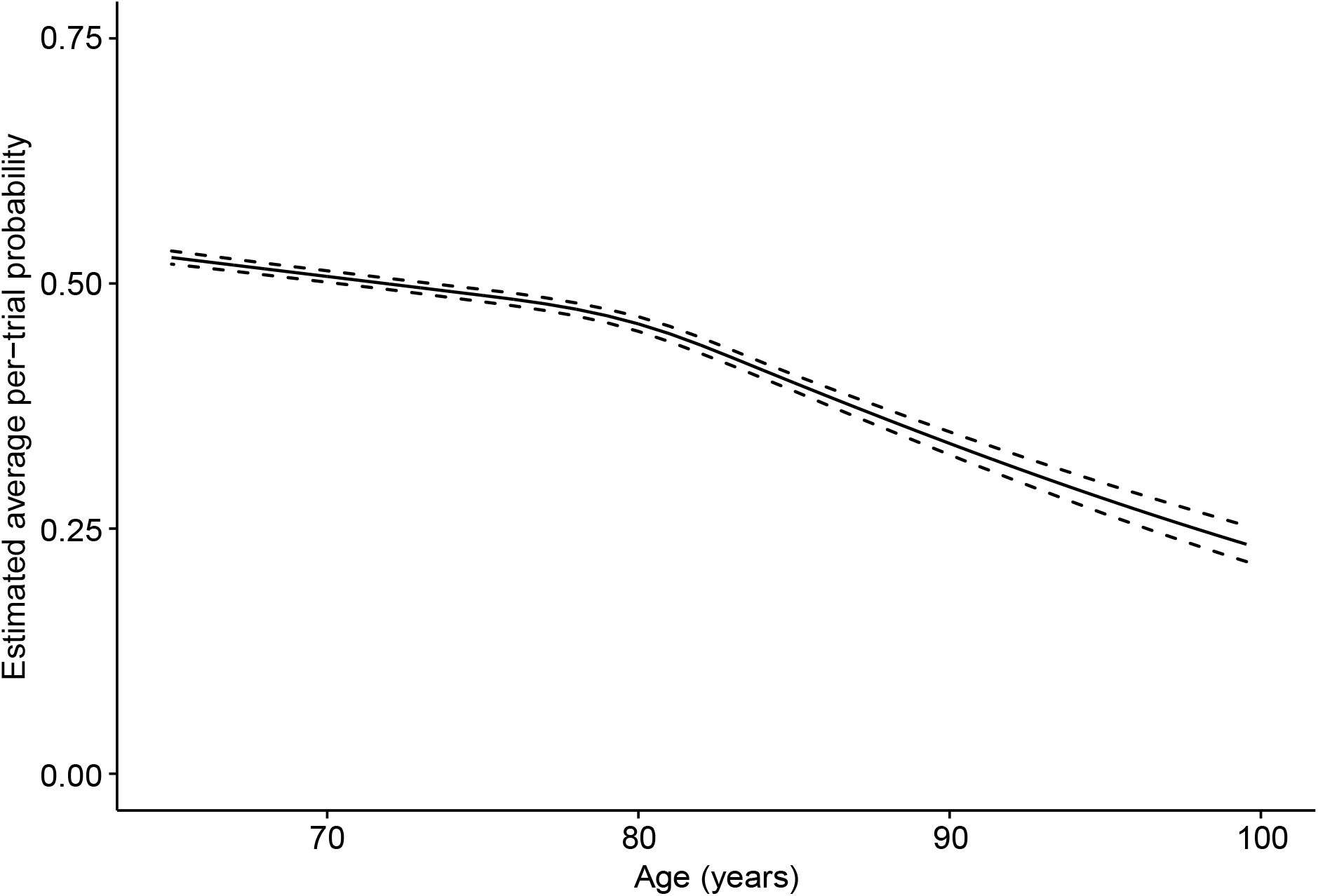
Predicted average per-trial probability (*p*) in the word recall test (with 95% Credible Interval) across age.

Figure 2 presents the estimated odds ratios (OR) for the remaining covariates in the linear predictor for *p*. It indicates the probability of correctly recalling words declined, on average, as the number of difficulties with ADLs increased, with an OR of 0.96 (95% Credible Interval 0.95, 0.97) indicating a 4% lower odds of recalling a word correctly for each activity reported to be performed with difficulty. Mean test score was lower when the interviewer recorded issues potentially affecting the test, with an OR of 0.77 (95% Credible Interval 0.75, 0.79). An estimated OR of 1.24 (95% Credible Interval 1.21, 1.27) indicated that the odds of recalling a word correctly were 24% higher for females than males. The odds were also higher in those with a higher level of education: compared to no educational qualifications, those with secondary educational qualifications had an OR of 1.31 (95% Credible Interval 1.28, 1.35), and those with HE qualifications had an OR of 1.54 (95% Credible Interval 1.50, 1.58). Finally, the odds were higher in later cohorts, with an OR of 1.16 (95% Credible Interval 1.14, 1.18) for each 1 S.D. (standard deviation) increase in the covariate (or 1.02 (95% Credible Interval 1.02, 1.02) for each year later participants were born).

**Figure 2.**
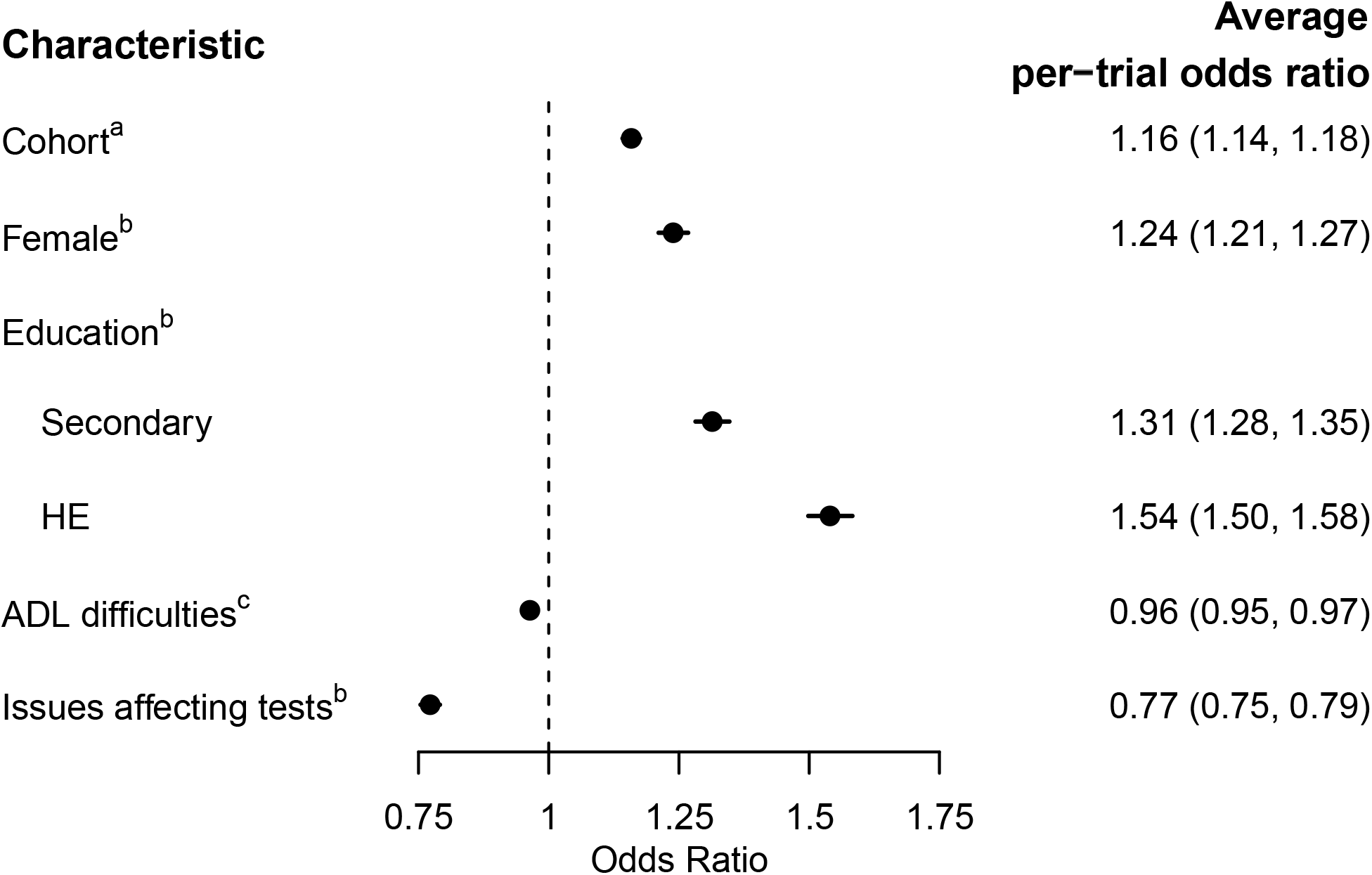
Estimated per-trial odds ratios (with 95% Credible Interval) for characteristics modelled in the linear predictor for average per-trial probability (*p*) in the word recall test. ^a^ The estimated average odds ratio for recalling a word correctly in the word recall test per 1 S.D. change in the predictor. ^b^ For categorical predictors: the estimated average odds ratio for recalling a word correctly in the word recall test when comparing the current category with the reference category. Reference categories as follows: Male (for Female); No qualifications (for Education); No issues which may have affected cognitive tests recorded (for Issues affecting tests). ^c^ The estimated average odds ratio for recalling a word correctly in the word recall test, for each additional difficulty reported with activities of daily living.

There was no evidence that the associations of any of these covariates with *p* differed across age (see Supplementary Materials).

### Association of covariates with *θ* (IIV)

Figure 3 presents the estimated change in the log of the intraindividual variability (IIV) or dispersion parameter, *θ*, for each modelled characteristic (NB lower values of *θ* indicate greater IIV: e.g. Supplementary Figure S1). It indicates that older people had greater IIV in memory scores, on average, with log(*θ*) estimated to be to be -1.38 (95% Credible Interval -1.70, -1.08) lower for each 1 S.D. increase in age at survey (or by -2.01 (95% Credible Interval -2.47, -1.57) for each decade of age). As Figure 3 also illustrates, higher educational qualifications were associated with lower IIV, with those who completed secondary school, for instance, having an estimated log(*θ*) higher by 0.54 (95% Credible Interval 0.18, 0.91), compared to individuals without qualifications. IIV was greater for individuals with more difficulties in their ADLs, with log(*θ*) reducing by an average of -0.25 (95% Credible Interval -0.35, -0.14) for each activity performed with difficulty. IIV was higher for females, with log(*θ*) lower by an average of -0.41 (95% Credible Interval -0.75, -0.09) compared to males. When the interviewer had recorded issues which might have affected the tests, the IIV was higher, with log(*θ*) reducing by an average of -2.76 (95% Credible Interval -3.13, -2.43). Later cohorts were also estimated to have greater IIV, with log(*θ*) lower by -1.01 (95% Credible Interval - 1.32, -0.73) for each 1 S.D. increase in the covariate (or -0.13 (95% Credible Interval -0.16, -0.09) for each year later participants were born).

**Figure 3.**
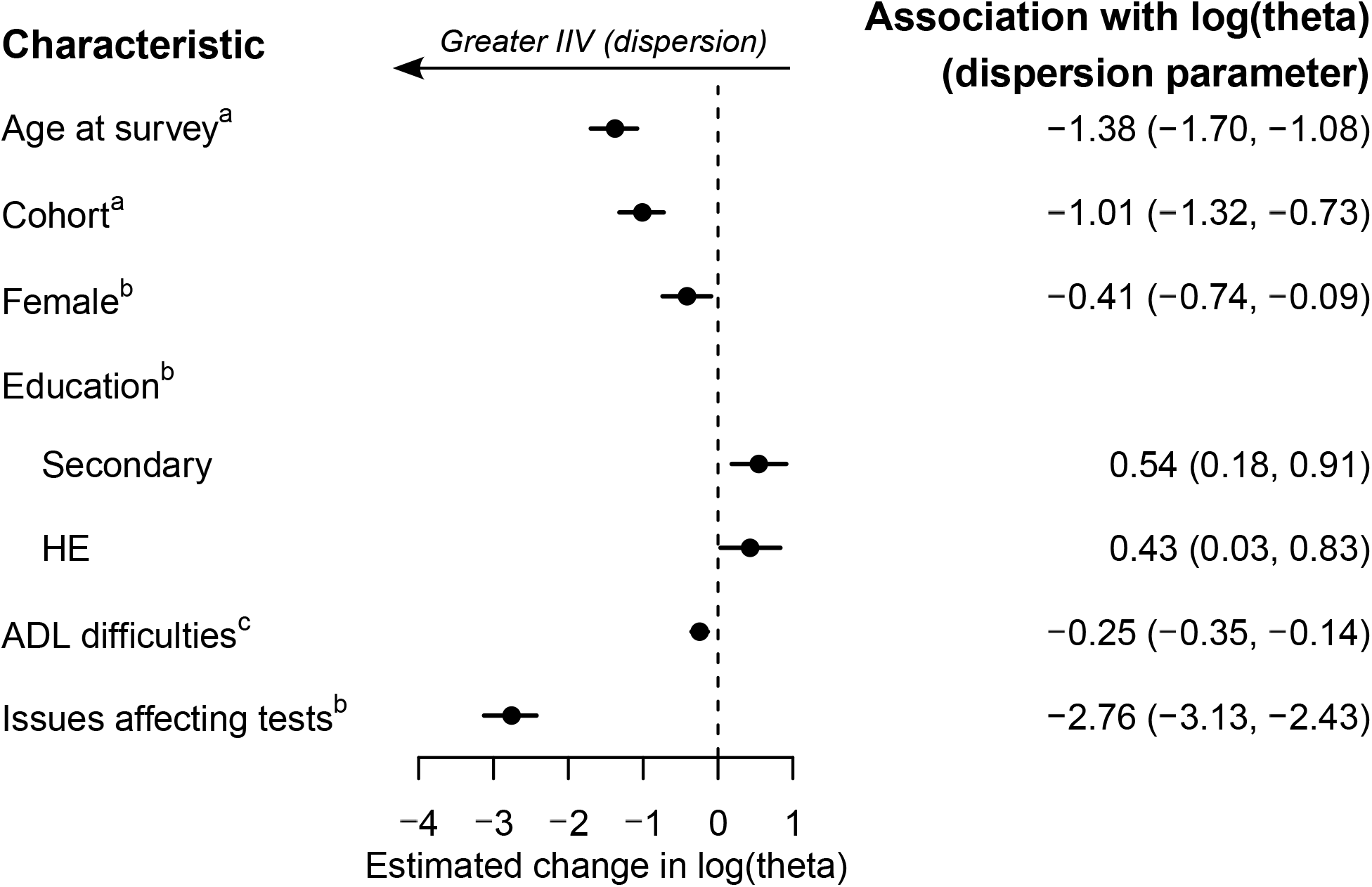
Estimated associations (with 95% Credible Interval) of modelled characteristics with IIV parameter (*θ*; theta) in the word recall test. Note that smaller values of log(*θ*) indicate greater dispersion (IIV). ^a^ The estimated change in log(*θ*) per 1 S.D. change in the predictor. ^b^ The estimated change in log(*θ*) when comparing the current category with the reference category. Reference categories as follows: Male (for Female); No qualifications (for Education); No issues which may have affected cognitive tests recorded (for Issues affecting tests). ^c^ The estimated change in log(*θ*), for each additional difficulty reported with activities of daily living.

### Participant effects

The random part of the model indicated that the correlation between average per-trial probability (*p*) at 70.5 years of age (random intercept) and rate of change in *p* (random slope) was estimated at 0.38 (95% Credible Interval 0.29, 0.48), suggesting that individuals with poorer episodic memory at age 70.5 years of age experienced a faster rate of decline in memory. Those who tended to score higher in the memory test (more positive random intercept for *p*) tended to have lower IIV (higher log(*θ*)), with a posterior mean correlation of 0.45 (95% Credible Interval 0.38, 0.53) between the random intercept (at 70.5 years of age) for *p* and the random intercept of the predictor for log(*θ*). Finally, the posterior mean correlation between the random slope for *p* and the random intercept for IIV (log(*θ*)) was 0.27 (95% Credible Interval 0.07, 0.48), indicating that those with slopes which decline less steeply (i.e. more positive estimates for the random slope) tended to have lower IIV (i.e. a higher estimate for log(*θ*)).

## DISCUSSION

We have shown that IIV in a visit-to-visit test of episodic memory in older English adults participating in the English Longitudinal Study of Ageing (ELSA) changed across age, sex, education, the number of difficulties with activities of daily living (ADLs), cohort, and reported challenges when performing the test. We simultaneously estimated IIV alongside the average per-trial probability in a mixed-effects location scale (MELS) beta-binomial model, finding that people with a higher probability of recalling words correctly tended to have lower IIV, and those with a more gradual decline in mean performance over time also tended to have lower IIV.

Our finding that IIV in the word recall test increased with age, whilst the probability of correctly recalling words decreased with age, on average, is characteristic of studies of cognitive functioning in advanced years, as reviewed in MacDonald *et al*., for example, who further discuss potential neural mechanisms underlying these age-related changes in IIV [10]. In addition, earlier analyses of the ELSA cohort have similarly found a faster average rate of decline in average word recall test performance at older ages [33, 34]. Higher mean memory performance in later cohorts, as we found, has also previously been reported in ELSA, and related to phenomena such as the Flynn effect [34], although we additionally found IIV to be greater in later cohorts too.

Our results also indicated that the greater the number of difficulties participants had with ADLs, the greater their IIV in memory test performance, on average, and the lower their probability of correctly recalling words too. The ability to perform ADLs is associated with cognitive, motor and perceptual functioning [35], and predicts mortality and morbidity [36, 37]: e.g. Fauth *et al*. found ADL disability predicted future dementia after controlling for baseline global cognitive status and other known risk factors [38]. Whilst we are not aware of any studies of IIV in cognitive performance and ADL functioning in older groups specifically, there have been studies of this association in other populations. For example, greater IIV as indexed by cognitive dispersion has been found to predict poorer functioning in basic ADLs in HIV-seropositive individuals without HIV-associated neurocognitive disorders [39].

We also found that lower educational levels were associated with greater IIV. In a study of within-occasion reaction time IIV in cognitive tests, Christensen *et al*. also found participants with fewer years of education had, on average, greater IIV [40]. Our results also indicated that, on average, lower educational levels predicted lower mean performance, mirroring the results of an earlier analysis of the same word recall test, using ELSA and the American Health and Retirement Study [41].

When interviewers indicated there were issues which may have affected the cognitive tests, then the probability of recalling words correctly was lower, on average, and IIV was greater. As Figure 3 indicates, this association was relatively large, and so test reliability is likely to be particularly low when such circumstances are reported, thus for researchers it is crucial to record any difficulties and take this into account in analyses.

We found people with a higher estimated probability of answering correctly at the sample mean age of 70.5 years (random intercept for *p*) had, on average, lower IIV (random intercept for log(*θ*)). Indeed, greater IIV in cognitive functioning has been previously found to typically predict lower mean scores [10]. This was also true at the population level for some of the covariates: for age, ADL functioning, education, and interviewer-recorded issues with cognitive tests, for instance, values of covariates which predicted higher IIV also tended to predict lower mean test performance. Sometimes, however, the converse was true: for example, whilst females were estimated to have higher mean memory score than males, their IIV was, on average, greater. With regard to the association of sex with mean performance, this concurs with Zaninotto *et al*., who also found females in the ELSA cohort to have a higher mean memory test score than males, with no moderating effect of age [33], although less explicit attention has been paid to the estimated effects of sex on IIV in memory-based tasks (*cf*. reaction time tasks, where females typically found to have greater IIV than males throughout adulthood) [42-44]. Similarly, whilst later cohorts were estimated to have a higher probability of correctly recalling words, their IIV was estimated to be greater too. This points to the utility of estimating the location (average per-trial probability, in our case) and scale (IIV) of repeatedly-measured cognitive tests: estimating quantities which are, to an extent, orthogonal, with each providing uniquely informative streams of information concerning underlying constructs of interest. We also found that those estimated to have poorer episodic memory at the sample mean age of 70.5 years experienced, on average, a faster rate of decline in memory, concurring with a latent group analysis of this outcome in ELSA participants aged 65-79 conducted by Olaya *et al*. [45]. In addition, our results indicated that those whose mean performance declined less steeply had, on average, lower IIV. At the individual-level, then, there tends to be a clustering together of higher mean performance, more gradual mean decline and lower IIV, and *vice versa*.

This study has several strengths. The ELSA cohort is designed to be representative of the older-aged English population, with comparisons of sociodemographic data with that from the national census suggesting this is broadly the case, and with further refreshment cohorts bolstering the representation of ages which attenuate with time [23]. By employing a MELS model, we were able to investigate the association of both time-varying and invariant factors with IIV, an opportunity lost if instead deriving an individual-level summary statistic for IIV. A MELS model also adjusts for sampling variability [18], unlike methods which do not allow the within-individual sample size to inform the estimate of IIV. In addition, by using a beta-binomial model we have applied an analytical method appropriate to bounded discrete outcomes [19], avoiding bias in estimates of IIV due to floor/ceiling effects [20].

This study also has limitations, however. There is the possibility of bias due to selection into the study. Whilst ELSA is designed to be representative of the target population, as in any longitudinal cohort there is attrition, and we additionally do not observe the outcome for people responding by proxy (since the word recall test cannot be administered by proxy). If selection into our analysis depends only on variables included in our models, such as age, sex, education, difficulties with ADLs, interviewer-recorded issues with cognitive tests, cohort, and observed values of the outcome, then our models will be unbiased. Having conditioned on these covariates, if selection into our analysis depends on the outcome, however, then there may be bias. Furthermore, we assume that the number of observations is independent of the underlying risk of drop-out (due to death, for example), which may otherwise lead to bias. Whilst there has been limited research into the issue of missing data in MELS models, it is an important issue starting to receive attention [46], with the need for further work, including for non-Gaussian outcomes, where the computational burden of methods designed to ameliorate bias is likely to be considerable [47]. In addition, our estimate of IIV depends on the fit of the model for the location (i.e. the linear predictor for average per-trial probability, *p*). The scientific significance of the modelled dispersion (IIV) parameter *θ* therefore depends on the choice of covariates included in the model for *p*, and also the extent of any measurement error they have (indeed, the same would be true of any analytical approach investigating residual error around a model for the location). We have chosen covariates which are, *a priori*, of interest, and have tried to keep a balance between parsimony and detail, but nevertheless the dispersion parameter, *θ*, may include variation which could be explained by a richer model for the location of the outcome.

In summary, analysing a memory test conducted with older people in the ELSA cohort, we found evidence of systematic differences in IIV, and also, having adjusted for those effects, evidence of residual between-individual differences in IIV. This indicates that sampling protocols for cognitive tests which rely on single, or just a few, measurement occasions to estimate mean groups differences can be prone to considerable measurement error [10]. At the population-level, IIV in cognitive functioning provided information which was orthogonal to mean performance, emphasising the importance of explicitly modelling IIV, rather than treating it as just a nuisance. In this study of visit-to-visit IIV, where measurements were made approximately every two years, inconsistency in task performance could be the result of both shorter (e.g. within-week, day, hour, etc.), and longer (e.g. over weeks or months), term changes in mean performance levels. Study designs which repeatedly-measure cognitive functioning over a variety of timeframes (repeated ‘bursts’ of measurement) would allow further characterisation of IIV over the shorter and longer term, which may differentially map onto underlying constructs of interest [48, 49]. Our study adds to the understanding of the factors associated with IIV in cognitive functioning in older ages, providing insights, beyond mean performance, into the biological mechanisms underlying differences in IIV, and their role in predicting future outcomes [6, 10, 12]. Future research is needed to investigate the impact of IIV on health and wellbeing.

## Supporting information

STROBE (EQUATOR network) checklist

## Data Availability

All data are available through the UK Data Service (SN 5050).

https://beta.ukdataservice.ac.uk/datacatalogue/studies/study?id=5050

## ACKNOWLEDGMENTS

The UK’s Medical Research Council (MRC) grant MR/N027485/1, awarded to investigator KT, funded RMAP. The MRC and the University of Bristol support the MRC Integrative Epidemiology Unit (MC_UU_00011/3). JKB was funded by MRC Unit Programme MC_UU_00002/5. The English Longitudinal Study of Ageing (ELSA) was developed by a team of researchers based at University College London, NatCen Social Research, the Institute for Fiscal Studies, the University of Manchester and the University of East Anglia. The data for ELSA were collected by NatCen Social Research. The funding for ELSA is currently provided by the National Institute on Aging in the US, and a consortium of UK government departments coordinated by the National Institute for Health Research. Funding for ELSA has also been received by the Economic and Social Research Council. The developers and funders of ELSA do not bear any responsibility for the analyses or interpretations presented here.

## DECLARATIONS

### Author contributions

Study concept and design: all authors; data preparation: RMAP; data analysis: RMAP; interpretation of results: all authors; drafting the manuscript: RMAP and GMZ; critical revision of the manuscript for important intellectual content: all authors. All authors have approved the final version of the manuscript. Authors GMZ and JKB share co-last authorship.

### Ethics approval

ELSA was conducted in accordance with the Declaration of Helsinki, and ethical approval was granted by the National Research Ethics Service (London Multicentre Research Ethics Committee (MREC/01/2/91)).

### Consent to participate

Participants gave their informed consent to take part in the study.

