## Supplementary Materials for "Intraindividual variability in cognitive function in older people in the English Longitudinal Study of Ageing: modelling risk factors using a mixed-effects beta-binomial model"

This supplementary material is from:

### EXAMPLE DISTRIBUTIONS FOR BETA-BINOMIAL PARAMETER VALUES

Figure S1 plots expected distributions of test scores for a beta-binomial model, given different values for the average per-trial probably parameter, *p*, and the intraindividual variability (IIV), or dispersion, parameter, $\theta$. E.g. assuming that a beta distribution, from which the probability underlying an observed count is sampled, has a *p* of 0.5, if this distribution is highly-dispersed (low $\theta$; e.g. $\theta = 0.5$), then the observed count is more likely to be distant from the mean than if the underlying beta distribution is less dispersed (e.g. $\theta = 4$) (Bolker, 2008; McElreath, 2020).

#### Figure S1. Mean probability of counts (from 20 trials) assuming beta distributions of underlying probabilities with different values of average per-trial probability (*p*) and dispersion (θ).

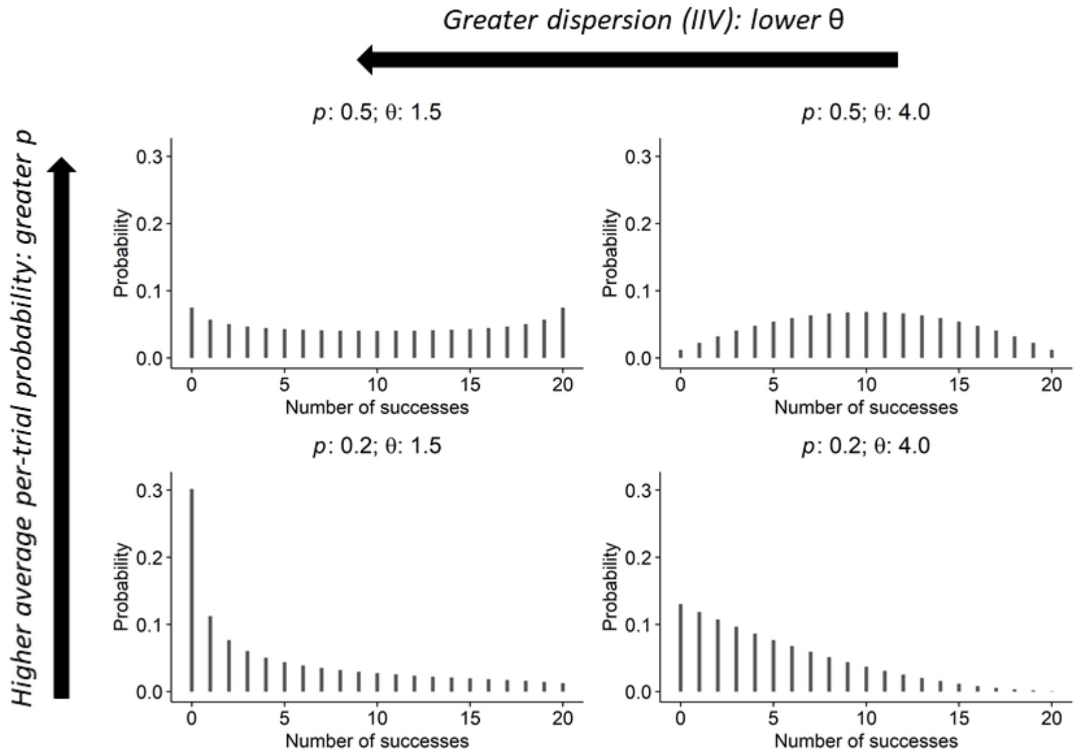

EXCLUSION & SELECTION

Figure S2. Flowchart detailing exclusions from final cohort (modelled dataset).

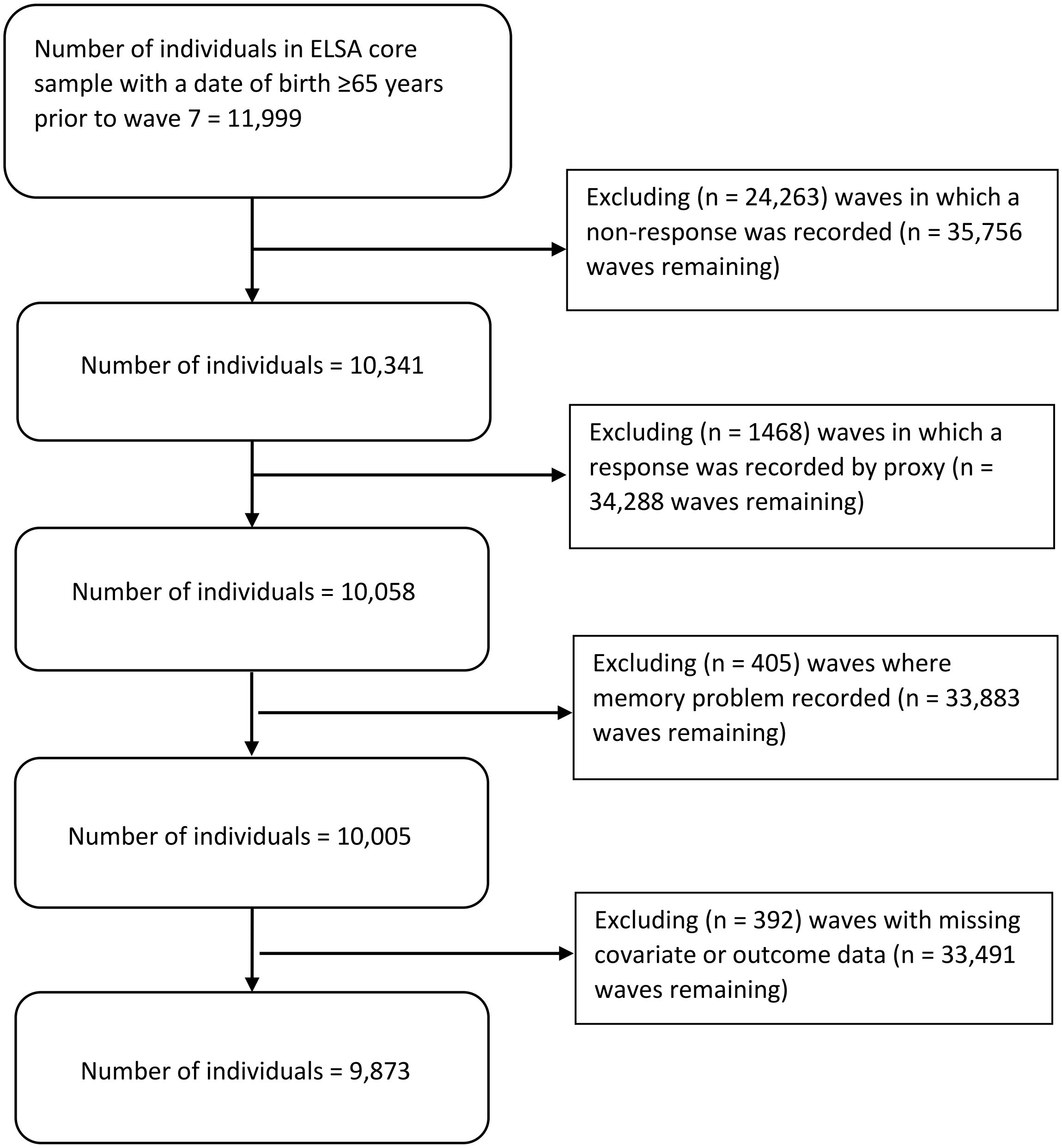

#### Table S1. Summary statistics for groups of participants subdivided by whether included in the final cohort (i.e. the analytical dataset) or not (all were ELSA core sample with a date of birth ≥65 years prior to wave 7).

| Mean (SD) or Number of individuals (%)[a] | Did not record a response for any eligible wave (n = 1,658) | Recorded a response for an eligible wave, but not included in model (n = 468)[b] | Included in model (n = 9,873) |
|---|---|---|---|
| Mortality | | | |
| Reported alive at wave 7 | 0 (0%) | 179 (38.2%) | 5194 (52.6%) |
| Reported dead during survey period | 296 (17.9%) | 155 (33.1%) | 2202 (22.3%) |
| Mortality status unknown at wave 7 | 1362 (82.1%) | 134 (28.6%) | 2477 (25.1%) |
| Interviewer recorded a memory problem in at least one wave[c] | N/A | | |
| Yes | | 94 (20.3%) | 396 (4.0%)[d] |
| No | | 369 (79.7%) | 9474 (96.0%)[d] |
| Unknown | | 5 | 3[d] |
| Sex | | | |
| Male | 814 (49.1%) | 246 (49.9%) | 1978 (44.4%) |
| Female | 844 (50.9%) | 222 (50.1%) | 2474 (55.6%) |
| Education at first survey | N/A | | |
| None | | 213 (58.0%) | 4731 (47.9%) |
| Secondary | | 82 (22.3%) | 2849 (28.9%) |
| Higher education | | 72 (19.%) | 2293 (23.2%) |
| Unknown | | 101 | 0 |
| Age at first survey | N/A | 71.8 (7.81) | 70.5 (6.4) |
| Cohort, year turned 65 | | | |
| 1971-1980 | 0 (0%) | 33 (7.1%) | 282 (2.9%) |
| 1981-1990 | 0 (0%) | 81 (17.3%) | 1595 (16.2%) |
| 1991-2000 | 0 (0%) | 93 (19.9%) | 3058 (31.0%) |
| 2001-2010 | 811 (48.9%) | 150 (32.1%) | 3574 (36.2%) |
| 2011-2020 | 847 (51.1%) | 111 (23.7%) | 1364 (13.8%) |

[a] Percentages based on valid N only.

[b] Not included in model due to: (a) the response being recorded by proxy, (b) the interviewer recording a memory problem, or (c) having missing data for any of the variables included in the model

[c] Memory problem defined as Alzheimer's disease or dementia, organic brain syndrome, senility or any other serious memory impairment. "No" / "Unknown" calculated from waves in which a response was recorded, only. If, instead, they are calculated from all waves in which the participant was eligible to respond, but did not necessarily do so (but was not recorded as dead) – in which case a smaller percentage of observations are made – then for those who recorded a response for an eligible wave, but not included in model, No: n = 166, Unknown: n = 208; and for those included in the model, No: n = 5897; Unknown: 3580.

[d] These are calculated from all waves in which the participant recorded a response, but not all responses were necessarily included in the model, hence the count for "Yes" and "Unknown" can be greater than 0. Surveys in which memory problems recorded were not included in model. When a memory problem was recorded, no subsequent waves for that participant were included in the model.

Table S2. Summary statistics for groups of participants subdivided by mortality status, and whether last survey included in model occurred in wave 7 or not.

| Characteristic at last survey included in model: Mean (SD) or Number of individuals (%) | Last observation prior to reported dead (n = 2,202)[a] | Last observation occurs prior to last wave (n = 4,956)[a] | Last observation is last wave (n = 4,917) |
|---|---|---|---|
| Age | 79.4 (7.6) | 77.0 (7.8) | 74.1 (6.9) |
| Cohort, year turned 65 | | | |
| 1971-1980 | 251 (11.4%) | 278 (5.6%) | 4 (0.1%) |
| 1981-1990 | 916 (41.6%) | 1410 (28.5%) | 185 (3.8%) |
| 1991-2000 | 811 (36.8%) | 1951 (39.4%) | 1107 (22.5%) |
| 2001-2010 | 224 (10.2%) | 1223 (24.7%) | 2351 (47.8%) |
| 2011-2020 | 0 (0%) | 94 (1.9%) | 1270 (25.8%) |
| Sex | | | |
| Male | 1108 (50.3%) | 2310 (46.6%) | 2180 (44.3%) |
| Female | 1094 (49.7%) | 2646 (53.4%) | 2737 (55.7%) |
| Education | | | |
| None | 1374 (62.4%) | 2901 (58.5%) | 1704 (34.7%) |
| Secondary | 492 (22.3%) | 1214 (24.5%) | 1682 (34.2%) |
| Higher education | 336 (15.3%) | 841 (17.0%) | 1531 (31.1%) |
| Number of activities of daily living performed with difficulty[b] | | | |
| 0 | 1250 (56.8%) | 3266 (65.9%) | 3953 (80.4%) |
| 1 | 440 (20.0%) | 801 (16.2%) | 506 (10.3%) |
| 2 | 233 (10.6%) | 420 (8.5%) | 247 (5.0%) |
| 3 | 133 (6.0%) | 237 (4.8%) | 117 (2.4%) |
| 4 | 106 (4.8%) | 169 (3.4%) | 69 (1.4%) |
| 5 | 40 (1.8%) | 63 (1.3%) | 25 (0.5%) |
| Issues with cognitive tests[c] | | | |
| No | 1679 (76.2%) | 3946 (79.6%) | 4532 (92.2%) |
| Yes | 523 (23.8%) | 1010 (20.4%) | 385 (7.8%) |

| | | | |
|---|---|---|---|
| Word recall test | 6.3 (3.7) | 7.1 (3.9) | 9.7 (3.7) |

[a] These are not mutually-exclusive categories.

[b] Sum of the activities of daily living (ADLs) where respondent reports any difficulty. ADLs include bathing, dressing, eating, getting in/out of bed, walking across a room.

[c] Interviewers recorded any issues which may have affected the participant's performance in the cognitive tests.

### MODEL BUILDING, ESTIMATION & SENSITIVITY ANALYSES

#### Model parameterisation

$$\text{Word Recall Test Score}_{ij} \sim \text{BetaBinomial}(N, p_{ij}, \theta_{ij})$$

$$\begin{aligned}
\text{logit}(p_{ij}) &= \beta_0 + \beta_1 \text{X}_{1ij} + \cdots + \beta_P \text{X}_{Pij} + u_{0j} + u_{1j}\text{Age}_{ij} \\
\log(\theta_{ij}) &= \alpha_0 + \alpha_1 \text{Age}_{ij} + \alpha_2 \text{Sex} + \alpha_3 \text{Cohort}_j + \alpha_4 \text{Sec Ed}_{ij} \\
&\quad + \alpha_5 \text{Higher Ed}_{ij} + \alpha_6 \text{ADL Difficulties}_{ij} + \alpha_7 \text{Test Issues}_{ij} + u_{2j}
\end{aligned}$$

$$\begin{pmatrix} u_0 \\ u_1 \\ u_2 \end{pmatrix} \sim \text{N}\left( \begin{pmatrix} 0 \\ 0 \\ 0 \end{pmatrix}, \Sigma_u \right)$$

$$\Sigma_u = \begin{pmatrix} \sigma_0^2 & & \\ \sigma_{01} & \sigma_1^2 & \\ \sigma_{02} & \sigma_{12} & \sigma_2^2 \end{pmatrix} = \begin{pmatrix} \sigma_0 & & \\ 0 & \sigma_1 & \\ 0 & 0 & \sigma_2 \end{pmatrix} \text{R} \begin{pmatrix} \sigma_0 & & \\ 0 & \sigma_1 & \\ 0 & 0 & \sigma_2 \end{pmatrix}, \quad \text{R} = \begin{pmatrix} 1 & & \\ \rho_{01} & 1 & \\ \rho_{02} & \rho_{12} & 1 \end{pmatrix}$$

**Equation 1**

Equation 1 depicts the mathematical formulation for the beta-binomial model used in our final analyses, with memory test score repeatedly-measured on occasion *i* (*i* = 1, …, $n_j$), within individual *j* (*j* = 1, …, J). *N* corresponds to the denominator of the test score (20). $\text{X}_{1ij}, \dots, \text{X}_{Pij}$ denote the *P* predictors added as fixed (population) effects for the average per-trial probability $p_{ij}$ (with logit link). These include age, as a non-linear function if appropriate, together with the other covariates as discussed in the main manuscript, and their interactions with the age term(s), again as appropriate. $u_{0j}$ and $u_{1j}$ denote the individual-level random effects for the intercept for $p_{ij}$, and also for the slope of age (as a linear term) for $p_{ij}$, respectively. The dispersion parameter $\theta_{ij}$ is allowed to depend on the fixed effects as indicated (i.e. those with coefficients $\alpha_1, \dots, \alpha_7$), and also on an individual-level random effect $u_{2j}$, with a log-link ensuring that it remains positive. The random effects are assumed to come from a multivariate Normal distribution, with unstructured covariance matrix $\Sigma_u$, and with corresponding standard deviations and correlations denoted *σ* and *ρ*, respectively (with subscripts reflecting the random effects they pertain to).

Note that there is an alternative parameterisation of the beta-binomial, with the beta distribution of the per-trial probability described by parameters *a* and *b*. When *a* = *b* = 1 then every per-trial probability between 0 and 1 is equally likely (describing a uniform distribution), whilst when *a* + *b* increases then the beta-binomial converges on the binomial distribution and the variance of the underlying heterogeneity decreases (Bolker, 2008). The parameterisation we used is often used in preference due to its more readily-interpretable nature, with *p* = average per-trial probability = *a* / (*a* + *b*) and *θ* = dispersion = *a* + *b.*

#### Model estimation

Bayesian estimation was via MCMC methods in Stan (2.21.0) (Stan Development Team, 2019), using the brms package (2.16.1) (Burkner, 2017), in R (4.1.0) (R Core Team, 2021).

Each model was fitted using four chains. Convergence to the target distribution was judged via visual diagnostics, the presence (and number and type) of any divergent transitions, and the value of split-$\hat{R}$

(with ≈ 1 suggesting convergence) (Gelman et al., 2013). The chains were run for such a length that (unless otherwise indicated) the bulk effective sample size (ESS) for each parameter of interest was a minimum of approximately 100 times the number of chains, indicating a reasonable number of independent draws from the posterior distribution. For models where random (individual) effects were saved (for leave-one-out cross validation, for example), thinning was applied so that the resulting chains were of a size more amenable to post-processing.

We used the priors specified by default by brms (Burkner, 2017). These included: an improper flat prior over the reals for the population-level (fixed) effects; a half student-t prior with 3 degrees of freedom and a scale parameter that depends on the standard deviation of the response after applying the link function for standard deviation of the random effects; an Lewandowski-Kurowicka-Joe (LKJ) prior with an eta of one for the correlations between the random (individual) effects; and a gamma(0.01, 0.01) prior for $\theta$ (used in some initial analyses for model selection where $\theta$ was assumed to be constant).

As sensitivity analyses we cross-checked evidence for the selection of fixed effects against models which assumed Gaussian residual error (but which otherwise had the equivalent fixed and random effects). Such models which assumed constant within-individual error (used when selecting a non-linear effect of age) were fitted using MLwiN (v.3.05), via R2MLwiN (v.0.8-7) in R (v.4.0.2) (Charlton et al., 2020; R Core Team, 2020; Zhang et al., 2016). When these were instead mixed-effects location scale Gaussian models (employed to cross-check selection of interaction terms), they were fitted using Bayesian estimation via MCMC methods in Stan (2.21.0) (Stan Development Team, 2019), using the brms package (2.16.1) (Burkner, 2017), in R (4.1.0) (R Core Team, 2021).

#### Comparing model fit

For the Bayesian models, model fit was assessed using Pareto smoothed importance sampling leave-one-out (PSIS-LOO) cross validation (CV) (Vehtari et al., 2017). PSIS is a method of approximating the average predictive accuracy of the training dataset (all the data bar one observation) for each observation left out (approximating the average predictive accuracy if we were to do this for each observation (fold), in turn, in the dataset). PSIS-LOO CV calculates the expected log predictive density (ELPD) as an estimate of out-of-sample predictive fit, where the model with the largest (most positive) ELPD has more support.

When some values of the estimate of the diagnostic shape parameter *k* of the generalised Pareto distribution (Pareto-$\hat{k}$) are > 0.7, then PSIS-LOO becomes less accurate, and K-fold CV is recommended as an alternative approach (Vehtari et al., 2017). K-fold differs from PSIS-LOO CV in that (a) the folds typically consist of many more than just one observation and (b) predictive accuracy is estimated from fitting many models (each to a dataset minus a particular fold) and then empirically testing predictive accuracy on the left-out fold (rather than estimating this predictive accuracy, as in PSIS-LOO). For the models in which we found (a small percentage of) values of Pareto-$\hat{k}$ to be > 0.7, then when possible we checked PSIS-LOO CV against K-fold CV, using K = 10 folds.

For the sensitivity analyses which assumed Gaussian residual error (see Model estimation) and constant within-individual error, we used the Akaike information criterion (AIC), with a reduction in AIC of ≥ 4 indicating the target model had more support (Burnham & Anderson, 2004) (for the sensitivity analyses which assumed Gaussian residual error but had a mixed-effects location scale (MELS) parameterisation, we instead used PSIS-LOO and K-fold CV, as described above).

#### Choice of non-linear function of age for the location of the outcome

Initial analyses were conducted to investigate the best-fitting function of age for the average per-trial probability parameter *p*. These were multilevel beta-binomial models, with measurement occasion (level 1) nested within participant (at level 2) (Steele, 2008). These models had a random intercept, and also a random slope for age (in decades) which was a linear term across its full range in the random part of the model, centred around its mean. They also included fixed effects for cohort and sex.

To investigate non-linear functions of age, we fitted restricted (aka natural) cubic splines of the covariate in the fixed part of the model for *p*. These consist of splines (sections of the regression line), between knots (pre-specified values of age where the splines are connected). The splines are cubic polynomials (although see below), with their first and second derivatives continuous at the knots, allowing the splines to smoothly-transition into each other. This property also allows them to be relatively insensitive to the location of knots, although the number of knots remains an important consideration: we used knot placements as recommended by Harrell, for models with k = 3 through to k = 7 knots (Harrell, 2015). 'Restricted' refers to the ends (before the first knot, and after the last knot) of the fitted line, which are restricted to be linear. In distributions with long tails, this reduces the influence of outliers.

We compared the fit of these models to each other, and also to a model with age as a linear term across its full range in the fixed part of the model for *p* (i.e. a model with no restricted cubic splines), using model fit statistics (see Model estimation and Comparing model fit, above) together with qualitative assessment of plotted predictions and posterior predictive checks (Gabry et al., 2019).

There was considerably more support for the beta-binomial models which allowed a non-linear function of age (via restricted cubic splines) compared to a model which instead fitted age as a linear term across its full range: for example, the model with k = 4 knots had the largest ELPD (in keeping with having most support), with the model with linear age having an ELPD -247.6 (SE: 20.2) lower. Otherwise, there was relatively little to choose between the restricted cubic spline models, with each having an ELPD slightly lower than the model with k = 4 knots, as follows: k = 6, ELPD -1.5 (SE: 7.8); k = 3 ELPD -7.3 (SE: 7.9); k = 5, ELPD -11.8 (SE: 7.8), with relatively large standard error for these differences. The model with k = 7 knots did not mix well, indicating this relatively elaborate parameterisation was not a practical one (a run of 4 chains, each with a warmup of 2000 iterations, and 2000 post-warmup iterations, took five days to run with chains in parallel, and yielded a minimum bulk ESS of 72, considerably lower than 100 times the number of chains (see Model estimation)).

A similar conclusion was drawn from the sensitivity analysis using the Gaussian models, which found the AIC was similar for k = 4 (AIC 166020.5), k = 5 (AIC 166018.4), k = 6 (AIC 166019.4) and k = 7 (AIC 166019.2) knots (although higher, indicating a poorer fit, for k = 3 (AIC 166055.4) knots, and considerably higher for the model in which age was linear across its full range (AIC 166218.1)). In these sensitivity analyses, the model with k = 4 was narrowly-favoured by a closed-test procedure (Royston & Sauerbrei, 2007).

There was therefore considerable evidence that the relationship between age and the location of the word recall test score was non-linear. The restricted cubic spline models all described a concave shape, with the rate of deterioration in mean word recall test score tending to increase with age, with close correspondence between the beta-binomial models and the sensitivity analyses using Gaussian models.

For the latter (Gaussian) models, this mean pattern was further confirmed using fractional polynomials (Royston et al., 1999), in which models with the lowest AIC described substantively the same pattern[1].

When choosing the number of knots in restricted cubic spline models, k = 4 or k = 5 provide a good choice for most datasets (Harrell, 2015). Qualitative assessment of plotted predictions, and posterior predictive checks, indicated close agreement between the beta-binomial models with k = 4 and k = 5 knots, with no substantive difference between these and predictions from more elaborate parameterisations. Given that k = 4 tended to come out equally (if not more) favourably than alternative parameterisations in the formal model selection exercises described above, and that it offers a good balance between a relatively parsimonious parameterisation on the one hand (when more complex functions may be more liable to present estimation issues when further elaborating the model) and sufficient flexibility to accommodate a range of patterns on the other (Harrell, 2015), a restricted cubic spline model with k = 4 knots was judged the best choice as a function for the fixed (population) effect of age for the average per-trial probability, *p*.

Once we had decided upon a non-linear function of age in the fixed part of the model for *p*, we also explored non-linear functions of age in the random part of the mean function, and also in the fixed part of the intraindividual variability function of the mixed-effects location scale models. However, given that (a) these models tended to have convergence problems, and (b) the relationship between the location of the word test score and age differed only modestly from a linear relationship, a more parsimonious model was favoured, in which age was included as a linear term across its full range in both the random part of the model for *p*, and in the fixed part of the model for intraindividual variability too.

#### Testing for interactions

To test for differences in the effect of covariates (sex, educational qualification, the number of ADLs with which the respondent reported difficulty, and whether the interviewer reported whether there were any factors which may have impaired the participants' performance during the cognitive tests) on average per-trial probability (*p*) across age, a beta-binomial model with main effects only was compared to models in which the age terms (linear term and two further spline terms) in the fixed part of the function for *p* were interacted with each of the covariates in turn.

Using PSIS-LOO cross validation (see Comparing model fit, above) this found that the standard error of the ELPD difference was considerably larger than 2 * the absolute value of the point estimate, indicating that there was no reliable evidence that the models were improved by fitting any of the interaction terms (sex interaction: ELPD diff -10.6 (SE diff 11.7); education interaction: ELPD diff 8.9 (SE diff 11.2); ADL interaction: ELPD diff -7.4 (SE diff 11.7); reported issues with cognitive tests interaction: ELPD diff -0.3 (SE diff 10.9)). Sensitivity analyses were conducted with Gaussian mixed-effects location scale models, in which the predictors were the same as in the beta-binomial case except now modelling the mean outcome on the original scale instead of *p*, and modelling log(within-individual SD) instead of

[1] In the fractional polynomial models, age was fitted as a transformed covariate. It was transformed by powers from the following set: -2, -1, -0.5, 0, 0.5, 1, 2, 3 (where 0 corresponds to the logarithmic transformation, and 1 corresponds to no transformation). We fitted a series of first-degree (FP1), and a series of second-degree (FP2), fractional polynomial models. For the FP1 models, only one power term is included for age in the model. Each of the powers from the set described above is used to transform age in a separate model, so eight models are fitted. For the FP2 models, two powers of the age are added to the model (with powers *p* and *q*: $\beta_0 + \beta_1 t^p + \beta_2 t^q$; NB if *p* = *q*, then $\beta_0 + \beta_1 t^p + \beta_2 t^p \log(t)$). We explored each power combination, so 36 models were fitted (44 models in total).

log(θ). This similarly found no reliable evidence that the models were improved by fitting any of the interaction terms in the function for the mean outcome (either via PSIS-LOO, nor via K-fold cross-validation using $k = 10$ folds, stratified on individual).

### MODEL ESTIMATES

#### Table S3. Results (untransformed estimates) from beta-binomial mixed-effects location scale model (from main manuscript).

| | | Mean | Lower 95% CI | Upper 95% CI |
|---|---|---|---|---|
| **linear model for average per-trial probability parameter (*p*): fixed (population-level) effects** | **Intercept** | -0.46 | -0.48 | -0.44 |
| | **Age (decades): linear[a]** | -0.16 | -0.18 | -0.13 |
| | **Age (decades): s1[a]** | -0.22 | -0.39 | -0.05 |
| | **Age (decades): s2[a]** | 0.09 | -0.45 | 0.62 |
| | **Cohort (year)[b]** | 0.02 | 0.02 | 0.02 |
| | **Female** | 0.21 | 0.19 | 0.24 |
| | **Highest educational qualification: secondary[c]** | 0.27 | 0.25 | 0.30 |
| | **Highest educational qualification: HE[c]** | 0.43 | 0.40 | 0.46 |
| | **No. activities of daily living with which have difficulty[d]** | -0.04 | -0.05 | -0.03 |
| | **Issues with cognitive tests** | -0.26 | -0.28 | -0.23 |
| **linear model for dispersion parameter (log($\theta$)): fixed (population-level) effects[e]** | **Intercept** | 7.93 | 7.27 | 8.68 |
| | **Age (decades): linear[a]** | -2.01 | -2.47 | -1.57 |
| | **Cohort[b]** | -0.13 | -0.16 | -0.09 |
| | **Female** | -0.41 | -0.74 | -0.09 |
| | **Highest educational qualification: secondary[c]** | 0.54 | 0.18 | 0.91 |
| | **Highest educational qualification: HE[c]** | 0.43 | 0.03 | 0.83 |
| | **No. activities of daily living with which have difficulty[d]** | -0.25 | -0.35 | -0.14 |
| | **Issues with cognitive tests** | -2.76 | -3.13 | -2.43 |
| **random (group-level) effects for individual[e]** | **SD(RE for intercept in function for *p*; u0)** | 0.46 | 0.45 | 0.48 |

| | | | | |
|---|---|---|---|---|
| | **SD(RE for age (decades) in function for *p*; u1)** | 0.21 | 0.17 | 0.24 |
| | **SD(RE for intercept in function for log($\theta$); u2)** | 2.85 | 2.56 | 3.18 |
| | **cor(u0, u1)** | 0.38 | 0.29 | 0.48 |
| | **cor(u0, u2)** | 0.45 | 0.38 | 0.53 |
| | **cor(u1, u2)** | 0.27 | 0.07 | 0.48 |

[a] Age at interview (decades). The linear term was centred around sample mean of 70.5 years (7.05 decades). Restricted cubic regression splines, with 4 knots (at the 5th, 35th, 65th and 95th percentiles) (Harrell, 2015) were fitted in the fixed part of the linear model for average per-trial probability (*p*); these constituted 3 terms (the linear term, and 2 further spline terms denoted s1 and s2). In the random part of the model for *p*, and in the fixed part of the model for the dispersion parameter ($\theta$), age was added as a linear term only.

[b] Cohort: year turned 65, centred around 1999.

[c] For highest educational qualification, the reference category is no educational qualification.

[d] Sum of the activities of daily living (ADLs) where respondent reports any difficulty. ADLs include bathing, dressing, eating, getting in/out of bed, walking across a room.

[e] when coefficients are estimated as *negative* in the linear model for the dispersion parameter (log($\theta$)), this implies that dispersion is greater (i.e. there is greater within-individual variability) with higher values of the associated covariate. (When $\theta = 2$, every probability equally likely; if $\theta > 2$, distribution of probabilities grows more concentrated; if $\theta < 2$ distribution is so dispersed that extreme probabilities near zero and 1 more likely than the mean).

### EXAMPLE MODEL CODE FOR MIXED-EFFECTS LOCATION SCALE BETA-BINOMIAL MODELS

Following Bürkner (Bürkner, 2021, 2022), we specified the mixed-effects location scale beta-binomial model by defining a custom response distribution using the brms library, in R.

```
library(“brms”)
# If wish to use all available cores to estimate model:
# options(mc.cores = parallel::detectCores())

# Define custom family for beta binomial
# for mu and phi (a.k.a. p and theta, respectively)):
beta_binomial2 <- custom_family(
  "beta_binomial2",
  dpars = c("mu", "phi"),
  links = c("logit", "log"),
  lb = c(NA, 0),
  type = "int",
  vars = "vint1[n]"
)

# Define stan functions:
stan_funs <- "
  real beta_binomial2_lpmf(int y, real mu, real phi, int T) {
    return beta_binomial_lpmf(y | T, mu * phi, (1 - mu) * phi);
  }
  int beta_binomial2_rng(real mu, real phi, int T) {
    return beta_binomial_rng(T, mu * phi, (1 - mu) * phi);
  }
"
stanvars <- stanvar(scode = stan_funs,
                    block = "functions")

# Specify formula, where: size = size of trial (20, in our case); person_ID
# is the group-level indicator (identifying each person, in our case);
# “s” (an arbitrary value, but one which importantly appears in each
# specification of the random part of the model) indicates we wish to allow
# for non-zero correlation between the random effects in each sub-model.
my_bf <- bf(
  outcome | vint(size) ~
    covariate_A + covariate_B +       # fixed effects for mu (a.k.a. p)
    (1 + covariate_A |s| person_ID),  # random effects for mu (a.k.a. p)
  phi ~ covariate_A + covariate_B +   # fixed effects for phi (a.k.a. theta)
    (1 |s| person_ID)                 # random effects for phi (a.k.a. theta)
  )

# Fit the model. NB: in case of an error evaluating the log probability at
# the initial value, it may help to set init_r (initial value) to a lower
# value than the (default) 2, e.g. init_r = 0.1 will set initial values as
# [-0.1, 0.1] rather than default [-2, 2]
# (see posts on https://discourse.mc-stan.org/ for further guidance)
brms_fit <- brm(
  formula = my_bf,
  data = df,
  family = beta_binomial2,
  stanvars = stanvars
```

```
)

# The following relates to post-processing models...

# expose self-defined Stan functions:
expose_functions(brms_fit, vectorize = TRUE)

# define log_lik functions (allowing e.g. leave-one-out cross-validation via
# loo() to be applied to model fit object):
log_lik_beta_binomial2 <- function(i, prep) {
  mu <- brms::get_dpar(prep, "mu", i = i)
  phi <- brms::get_dpar(prep, "phi", i = i)
  trials <- prep$data$vint1[i]
  y <- prep$data$Y[i]
  beta_binomial2_lpmf(y, mu, phi, trials)
}

# define posterior_predict method (allowing e.g. posterior predictive checks
# via pp_check() to be applied to model fit object):
posterior_predict_beta_binomial2 <- function(i, prep, ...) {
  mu <- brms::get_dpar(prep, "mu", i = i)
  phi <- brms::get_dpar(prep, "phi", i = i)
  trials <- prep$data$vint1[i]
  beta_binomial2_rng(mu, phi, trials)
}
```

See Bürkner for further guidance (Bürkner, 2021, 2022).
